# Development of molecular subtype specific prognostic marker signature in immune response associated colon cancer through fuzzy based transcriptomic approach

**DOI:** 10.1101/2023.05.18.23290045

**Authors:** Necla Koçhan, Barış Emre Dayanç

**Author notes:** To whom correspondence should be addressed Barış Emre Dayanç; Address: Izmir University of Economics, Balcova, Izmir, Turkey; Tel: (+90).

## Abstract

**Objective:** The molecular heterogeneity of colon cancer makes the prediction of disease prognosis challenging. In order to resolve this heterogeneity, molecular tumor subtyping present solutions. These approaches are expected to contribute to clinical decision-making. In this study, we aimed to identify Consensus Molecular Subtype (CMS) specific prognostic genes of colon cancer, focusing on anti-tumor immune-response associated CMS1, through a fuzzy-based machine learning approach.

**Materials and Methods:** We applied Fuzzy C-Means (FCM) clustering to stratify patients into two groups and identified genes that predict significant disease-specific survival difference between groups. We then performed Cox regression analyses to identify the most significant genes associated with disease-specific survival. A subtype-specific risk score and a final risk score formulae were constructed and used to calculate risk scores to stratify patients into low and high-risk groups within each CMS (1 to 4) or independent of CMS respectively.

**Results:** We identified CMS-specific genes and an overall 11-gene signature for prognostic risk prediction based on the disease-specific survival of colon cancer patients. The patients in both discovery and test cohorts were stratified into high and low-risk groups using subtype risk scores. The disease-specific survival of these risk groups within each CMS, except CMS3, was significantly different for both discovery and test cohorts.

**Discussion and Conclusions:** We have identified novel prognostic genes with potential immune regulatory roles within the immune-response associated CMS1. The low number of patients in the CMS3 cohort prevented subtype-specific prognostic gene validation. Tumor stage grouping of the validation cohort suggested the best prediction of prognosis in tumor stage III patients. In conclusion, newly identified eleven genes can efficiently predict the prognostic risk of colon cancer patients and classify patients into corresponding risk groups.

**Graphical Abstract:** 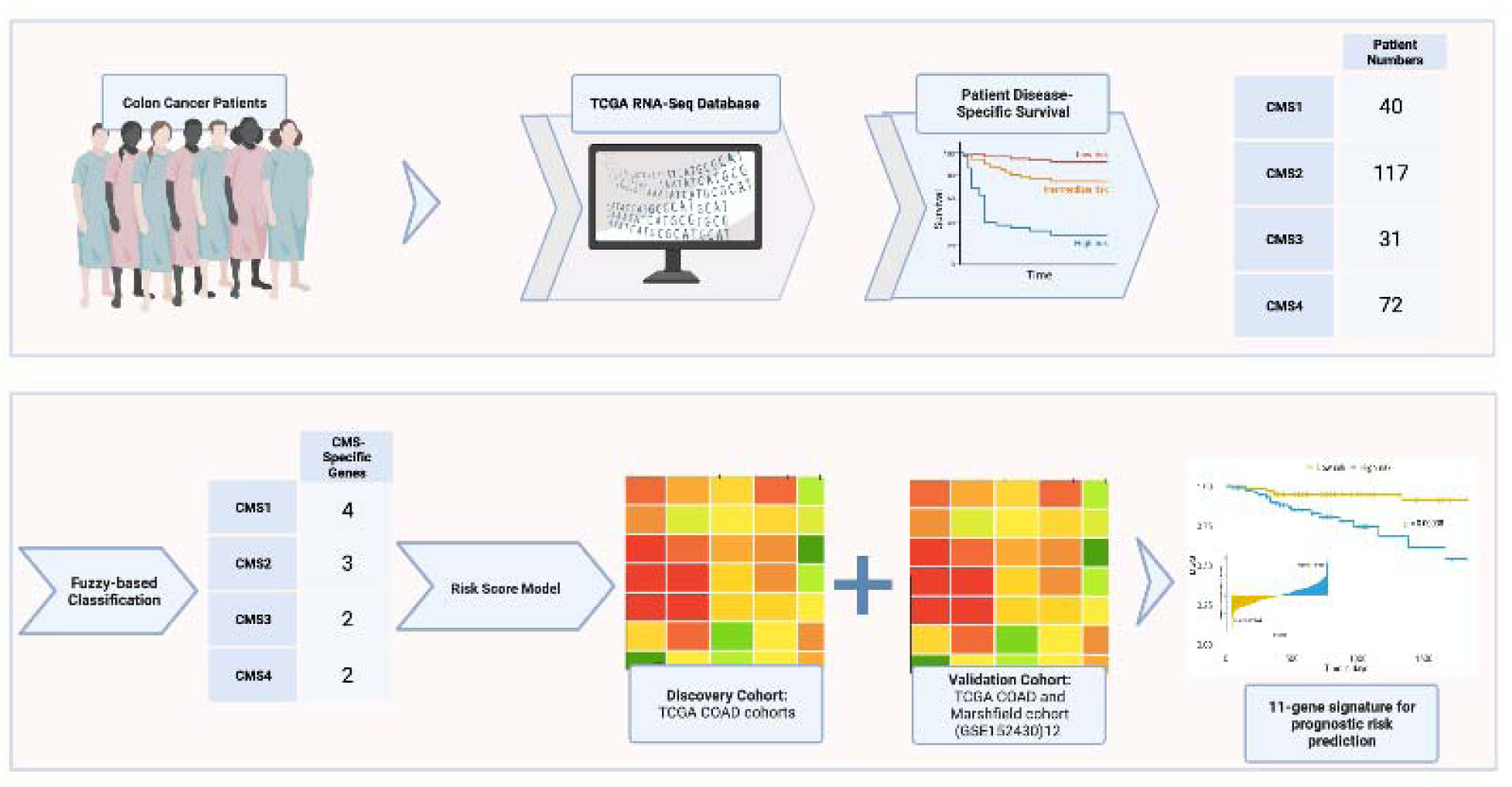

## BACKGROUND AND SIGNIFICANCE

Colon cancer is a disease with a low survival rate^1^. It is one of the most common cancer types worldwide and it ranks 4^th^ in the world in terms of incidence with 1,148,515 new diagnoses in 2020, and 576,858 deaths (about 6% of cancer-related deaths) reported worldwide in the same year according to global cancer statistics^2^. Epidemiological studies suggest that increased colon cancer incidence is associated with advanced age and smoking status, decreased physical activity, diet and obesity^3^. Loss of genomic and/or epigenomic stability is observed in the formation of neoplastic lesions associated with the onset of colon cancer. Earlier studies suggested that this loss of stability leads to cancer by increasing the accumulation of mutations in oncogenes and tumor suppressor genes.^3^

Colon cancer diagnosis is associated with patient’s symptoms such as blood in the stool, changes in bowel habits and abdominal pain. Other symptoms include fatigue, pallor, shortness of breath, weight loss and anemia^3^. Colonoscopy is still the gold standard for the diagnosis of the disease, allowing biopsies during imaging and further histological and pathological examination leading to some molecular profiling with diagnosis. Colonoscopy is also used for the monitoring of the disease during therapy. While the most common diagnostic method for colon cancer is colonoscopy, there are several methods currently in use and in development such as capsule endoscopy, CT colonoscopy, markers, endoscopy and sigmoidoscopy ^3^. The gold standard for monitoring the prognosis in colon cancer is clinicopathological staging (TNM or AJCC classification). Emerging -*omics technologies* are expected to provide alternative approaches for predicting and monitoring colon cancer prognosis.

Microarray and next generation sequencing (NGS) are widely used omics technologies, which enable researchers to analyze the molecular characteristics of cancer and reveal more detailed information about the complex structure of cancer at gene expression level. The data obtained through these new technologies have been used to identify the genes that affect the survival of colon cancer patients previously ^4–8^. Currently both RNA-sequencing (RNA-seq) and microarray data were used in research studies to validate results. In our studies, we preferred RNA-seq data for our analyses.

Previously, colon cancer classification identified that approximately 17% of colon cancer patients exhibit microsatellite instability (MSI), 60% have chromosomal instability (CIN) and 20% exhibit CpG island methylation phenotype (CIMP), hence classified as CIN^+^, MSI^+^, CIMP^+^ or triple negative cancers. Recently, colon cancer began to be classified within four consensus molecular subtypes (CMSs) and unclassified samples^1^. For each CMS, biological characteristics (e.g. immune-response association for CMS1) and associated mutations (e.g. KRAS mutation for CMS3) were identified. In summary, CMS1 is defined by robust immune response, microsatellite instability (MSI), hypermutation, hypermethylation (associated with CIMP), low somatic copy number changes, high BRAF mutations and overexpression of DNA mismatch repair (MMR) molecules. For CMS2, epithelial differentiation, excessive WNT and MYC signal activation has been observed with high chromosomal instability (CIN) resulting in frequent copy number gains in oncogenes and copy number losses in tumor suppressor genes. CMS3’s defining characteristics include KRAS-activated mutations resulting in significant metabolic disorder, high CIN and increased oncogene expressions. Finally, CMS4 shows activation of genes involved in the epithelial-to-mesenchymal transition (EMT), high CIN and significant TGF-β activation. Unclassified tumors were defined as any samples that do not show sufficient overlap with described subtypes with consistent characteristics.^1^

With the recent proposal of four transcriptomic-based molecular subtypes, the value of using molecular subtyping for prognostication of colorectal cancer became further evident ^9–10^. Bramsen *et al.* (2017) concluded that subtype-specific genes improve the prediction of prognosis in colorectal cancer. Additionally, Purcell *et al.* (2019) points out that the absence of targeted therapy for primary colorectal cancer suggested the value of tumor stratification with CMSs as a prognostic approach. These studies provided us the rationale for focusing on the identification of consensus molecular subtype-specific genes through a fuzzy-based approach, where fuzzy handles the uncertainty and ambiguity in the data and assigns samples to the clusters with respect to the degree of membership. For this purpose, we first employ Fuzzy C-Means (FCM) clustering in order to stratify colon cancer patients regarding gene expressions. Then for each gene, we apply Kaplan-Meier (KM) analysis with log-rank tests in order to identify genes that can significantly differentiate between the patient’s survival within each CMS. Next, Cox regression analyses were performed to calculate the subtype specific risk scores and predict prognosis.

## MATERIALS AND METHODS

### Gene expression data and survival data

RNA-Seq data for CRC patients are obtained from the TCGA RNA-Seq database using the TCGAbiolinks package. We focused on patients with Primary Tumor (PT) and accompanying Solid Tissue Normal (STN). Among these patients, our cohort was limited to patients with colon adenocarcinoma diagnosis but received no therapy. We used 5-year Disease Specific Survival (DSS) data for the survival analysis, as DSS is more stringent than Overall Survival (OS) in colorectal cancer.^11^ The DSS data of TCGA COAD samples is kindly provided by the authors ^7^. Since our aim is to focus on the molecular subtype specific prognostic genes in CRC, we used CMS information of TCGA COAD patients from the Synapse website (https://www.synapse.org/). Patient “TCGA-CK-6746” was excluded from the TCGA COAD data due to unspecified DSS. Our study cohort consisted of 40 patients in CMS1, 117 patients in CMS2, 31 patients in CMS3 and 72 patients in CMS4. 77 percent of the patient data formed the discovery set among the CMSs. We filtered the genes below 0.5 Fragments Per Kilobase of transcript per Million (FPKM) in both PT and STN to avoid systematic bias of RNA-Seq data on genes with low expression. After filtering, 14,334 genes were used for downstream analysis. For the validation cohort, we used TCGA COAD and Marshfield cohorts (GSE 152430)^12^ with available RNA-seq and DSS data in primary colon tumors.

### Identification of subtype specific prognostic genes for colon cancer using FCM

In order to identify subtype specific prognostic genes, analyses were performed for each CRC molecular subtype separately. The FCM clustering algorithm was applied to stratify patients into two clusters (groups) with membership degrees for each patient in each cluster. The algorithm assigns each patient to one of the clusters with the maximum membership degree, which displays the degree of belonging to the corresponding cluster. A representative violin plot for CXCL10 gene out of 14,334 genes was shown in Supplementary Figure S1.

### Risk score model

For any subtype s, we define subtype specific risk score for a patient, denoted by *R_s_*, as follows:

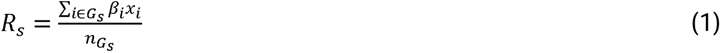

where *G_s_* is the set of genes in the signature gene list of the subtype *s*, *n_Gs_* is the number of genes in *G_s_*, *β_i_* and *x_i_* are Cox coefficient and the expression value of gene *i*, respectively. Each subtype risk score is scaled using the “scale” function in R, which normalizes the risk score values. Then final risk score of a patient, denoted by *R*, is defined as weighted combination of the subtype specific risk scores:

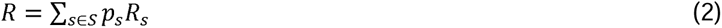

where *p_s_* is the probability of a patient belonging to the subtype s such that ∑*_sES_ p_s_* = 1. Since the sum of the probabilities is 1, the final risk score will have the same scale as subtype specific risk scores. We then divide samples into two groups (high-risk vs low-risk) using mean cutoff of the final risk score.

### Statistical analysis

Statistical analyses were performed using R language (v.4.0.2). Kaplan-Meier and log-rank test were performed to assess survival differences between clusters and risk groups. Univariate and multivariate Cox regression analyses were performed using “survival” and “survminer” packages in R. A stepwise variable selection procedure (“My.stepwise’’ package from R) was applied to obtain the best candidate Cox proportional hazards model and so the most informative genes. The genes leading to convergency problem in “My.stepwise.coxph” function were eliminated. *p* values below 0.01 and 0.05 were considered statistically significant for all comparisons and labelled as such.

## RESULTS

In order to identify subtype specific genes for colon cancer patients, we followed the process given in Figure 1. After preprocessing and splitting the data based on the CMSs, we first applied Fuzzy C-Means (FCM) clustering in order to stratify patients into two groups. In Supplementary Figure S1, we showed that the normalized expression values (log_2_+1) of representative gene CXCL10 out of 14,334 genes for each consensus molecular subtype demonstrated stratification into two groups according to the membership degrees of each patient belonging to the corresponding clusters.

**Figure 1.**
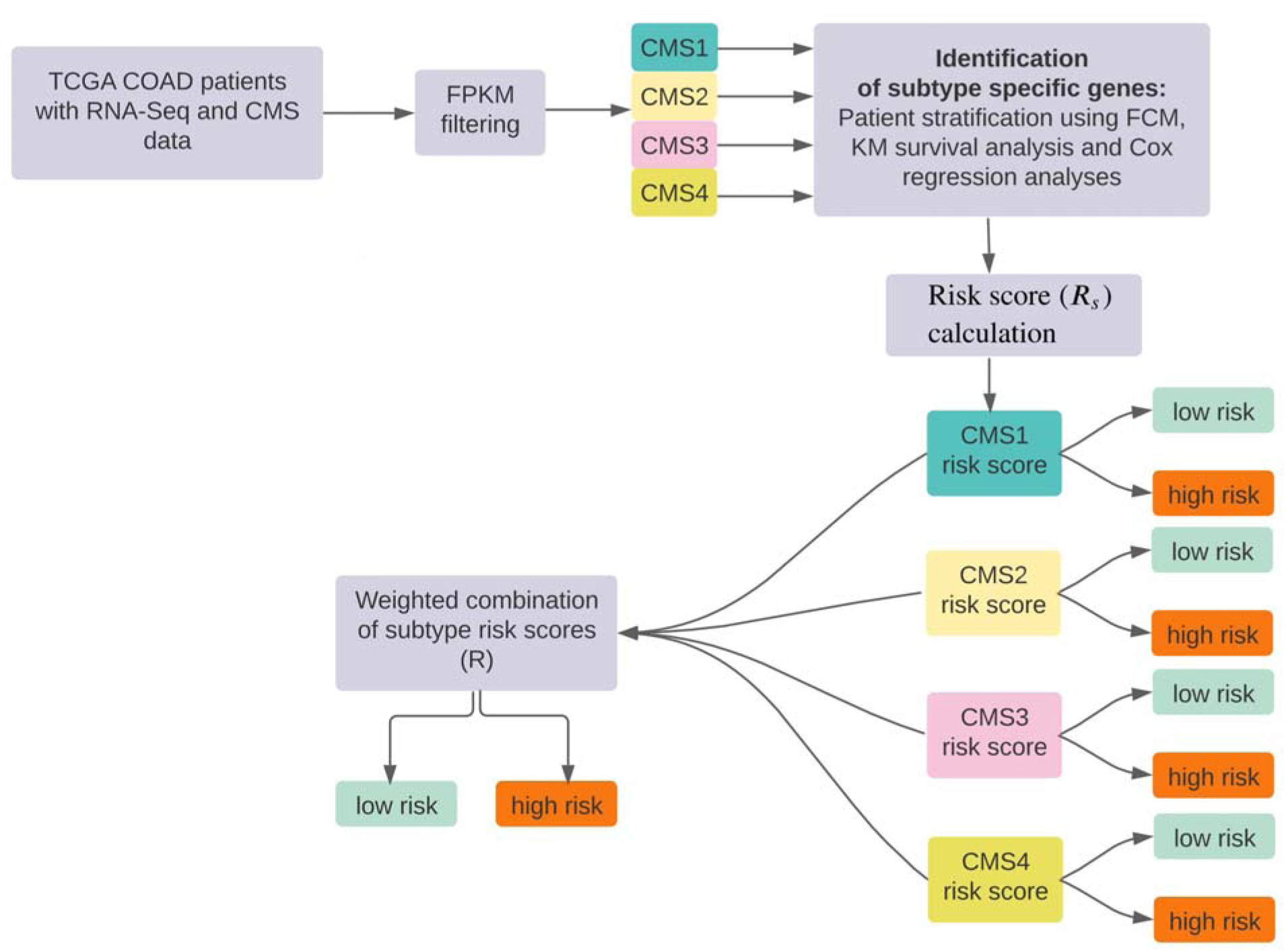
Flow chart of the proposed method in order to identify subtype-specific genes for RNA-Seq TCGA COAD patients.

For each subtype, we performed KM survival analysis with log-rank test to identify genes that can significantly differentiate between survival times of the subgroups obtained from FCM clustering with a cutoff *p* value of 0.001. We obtained 72 genes for CMS1, 69 genes for CMS2, 8 genes for CMS3 and 253 genes for CMS4 that are statistically significant (*p<*0.001). We then performed univariate and multivariate Cox regression analyzes to identify the significant genes that are predictive of survival. We note here that since we could not find any significant genes for CMS3 when cutoff value of p was considered 0.01, we have taken *p*-value as 0.05 only for CMS3. As a result, we found four genes for CMS1, three genes for CMS2, two genes for CMS3 and two genes for CMS4, listed in Table 1. We also calculated the FDR adjusted *p*-values of subtype specific genes within each CMS. Cox coefficients, *p*-values, hazard ratios and False Discovery Rates (FDR) of each of 11 signature genes were listed in Table 1.

**Table 1.** Prognostic values of 11 genes from TCGA-COAD training set.

| CMS | Symbol | NCBI<br>Entrez<br>Gene ID | Description | Cox coeff. | $p$-value | HR (95% CI) | FDR |
| --- | --- | --- | --- | --- | --- | --- | --- |
| **CMS1** | VSTM5 | 387804 | V-set and transmembrane domain containing 5 | -0.6533 | 0.0072 | 0.5203 (0.3231-0.8377) | 0.0088 |
|  | ZBTB7C | 201501 | Zinc finger and BTB domain containing 7C | -0.7075 | 0.0069 | 0.4929 (0.2950-0.8235) | 0.0088 |
|  | BCAS1 | 8537 | Breast carcinoma amplified sequence 1 | -0.7334 | 0.0070 | 0.4803 (0.2820-0.8181) | 0.0088 |
|  | CXCL10 | 3627 | C-X-C motif chemokine ligand 10 | 0.7945 | 0.0088 | 2.2133 (1.2216-4.0100) | 0.0088 |
| **CMS2** | FOXJ1 | 2302 | Forkhead box J1 | -0.3499 | 0.0079 | 0.7048 (0.5444-0.9125) | 0.0079 |
|  | PCDHB16 | 57717 | Protocadherin beta 16 | 0.5776 | 0.0036 | 1.7818 (1.2072-0.7551) | 0.0079 |
|  | RNF125 | 54941 | Ring finger protein 125 | -0.9937 | 0.0063 | 0.3702 (0.1815-0.7551) | 0.0079 |
| **CMS3** | R3HCC1 | 203069 | R3H domain and coiled-coil containing 1 | -1.9572 | 0.0387 | 0.1413 (0.0221-0.9033) | 0.0468 |
|  | NKX6-3 | 157848 | NK6 homeobox 3 | 0.3170 | 0.0468 | 1.3731 (1.0045-1.8769) | 0.0468 |
| **CMS4** | SLC29A4 | 222962 | Solute carrier family 29 member 4 | 1.0155 | <0.001 | 2.7606 (1.5283-4.9865) | <0.001 |
|  | RHCG | 51458 | Rh family C glycoprotein | -0.7483 | 0.0021 | 0.4732 (0.2939-0.7617) | 0.0021 |

Abbreviations: HR, Hazard Ratio; CI, Confidence Interval; FDR, False Discovery Rate.

To evaluate the subtype risk score for each colon cancer patient, we applied the risk score formula previously defined in (1) using Cox coefficients given in Table 1. We then divided samples into two groups (high-risk vs low-risk) using mean cutoff of the subtype risk scores. The KM survival analysis displayed that the patients with high-risk scores in each subtype showed significantly poorer prognosis than those with low-risk scores in the discovery cohort (Figure 2, *p<0.05*).

**Figure 2.**
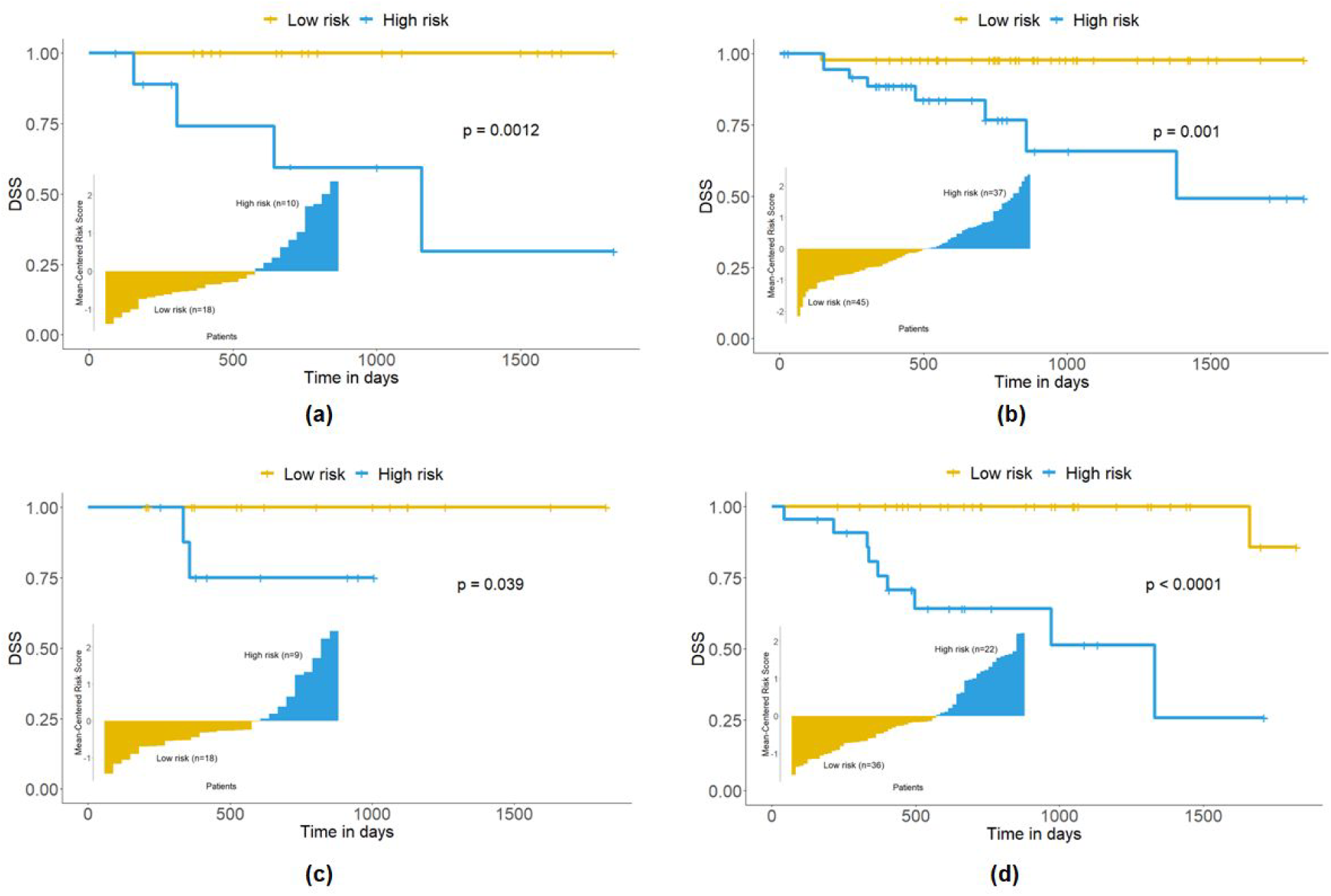
Patient stratification based on subtype risk scores with CMS specific prognostic markers. Bar charts and KM survival curves for CSM1 (a), CMS2 (b), CMS3 (c) and CMS4 (d). Risk scores are given in a bar chart, where each bar indicates the subtype risk score for an individual colon cancer patient. KM curve showing colon cancer patients with high and low-risk groups. Subtype-specific signature genes can distinguish high-risk group from low-risk group within each subtype.

Our approach can also stratify the patients into two groups using these 11 genes regardless of molecular subtypes, into low-risk group (102 patients) and high-risk group (93 patients). Kaplan-Meier analysis with log-rank test showed that our risk score approach can distinguish low-risk group from high-risk group given these subtype-specific signature genes (Figure 3, *p*<0.001). Like AJCC staging, our risk model can significantly predict the prognosis of colon cancer patients. However, we obtain a higher hazard ratio when our risk model is applied. Results were summarized below in Table 2.

**Figure 3.**
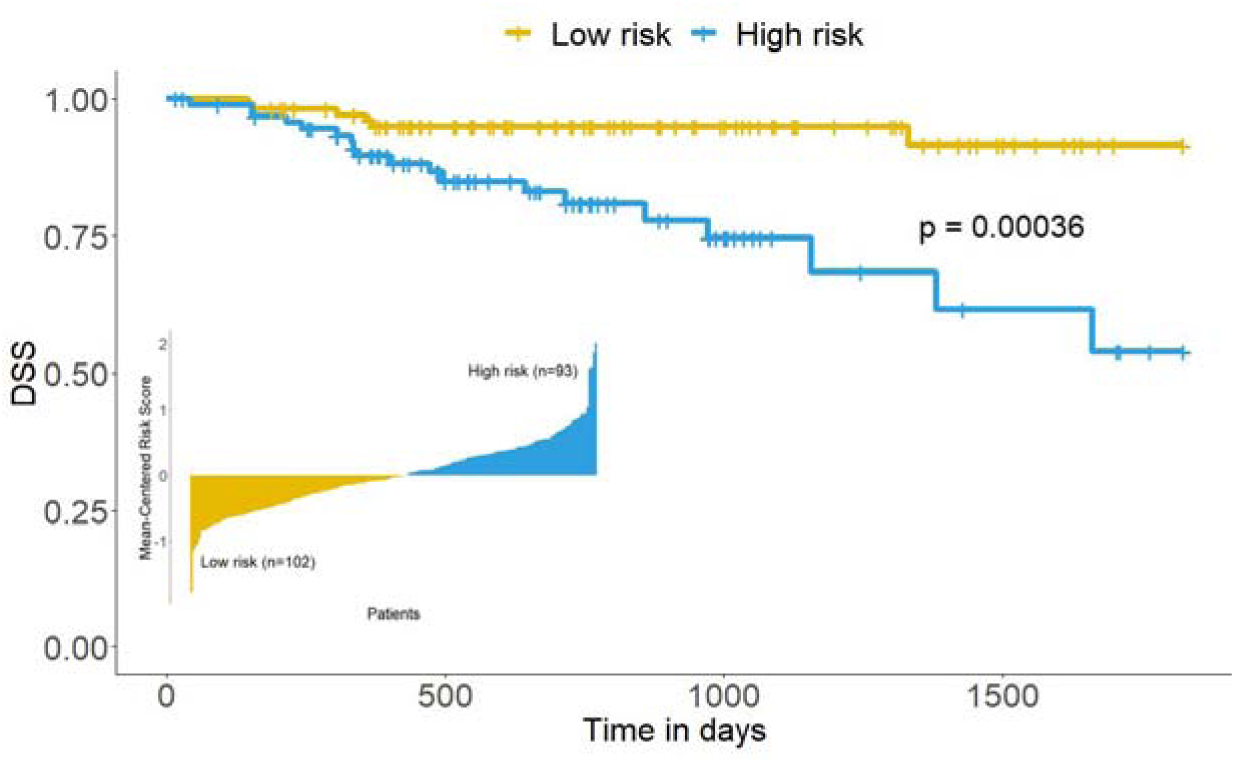
CRC patient stratification with 11 prognostic markers. Risk scores are given in a bar chart, where each bar indicates the final risk score for an individual colon cancer patient. KM curves showing colon cancer patients with high and low-risk groups. Subtype-specific signature genes can distinguish high-risk group from low-risk group with the log-rank *p*-value of 0.00036. Survival times are in days.

**Table 2.** Univariate and multivariate Cox regression analysis of clinical parameters in discovery and validation cohorts.

|  | Variable | Univariate |  |  | Multivariate |  |  |
| --- | --- | --- | --- | --- | --- | --- | --- |
|  |  | HR | 95% CI | p-value | HR | 95% CI | p-value |
| Discovery cohort | Gender (female vs. male) | 0.75 | 0.353-1.591 | 0.452 | 1.22 | 0.51-2.90 | 0.655 |
| | Age at diagnosis (< 65 vs. ≥ 65) | 1.24 | 0.582-2.65 | 0.577 | 2.48 | 1.01-6.12 | 0.048 |
|  | Microsatellite status<sup>+</sup> | 1.01 | 0.36-2.80 | 0.990 |  |  |  |
| | CEA level (< 28 vs. ≥ 28) | 1.00 | 0.99-1.02 | 0.457 | | | |
|  | AJCC stage<sup>++</sup> | **3.24** | **1.97-5.32** | **<0.001\*\*** | **3.38** | **2.01-5.71** | **<0.001\*\*** |
|  | **Risk model (low-risk vs. high-risk)** | **4.66** | **1.85-11.76** | **0.001\*\*** | **3.95** | **1.60-9.75** | **0.003\*** |
| Validation cohort | AJCC stage<sup>++</sup> | **3.60** | **2.37-5.46** | **<0.001\*\*** | **3.70** | **2.43-5.64** | **<0.001\*\*** |
|  | **Risk model (low-risk vs. high-risk)** | **2.84** | **1.33-6.07** | **0.007\*** | **2.81** | **1.30-6.07** | **0.009\*** |

Significance: less than 0.001 \*\*\*, less than 0.01 \*\*.
+ Microsatellite status: Treated as continuous variable (1: MSI-L, 2: MSI-H, 3: MSS)
++ AJCC stage: Treated as continuous variable (1: stage I and IA, 2: stage II, IIA, IIB and IIC, 3: stage III, IIIA, IIIB and IIIC, 4: stage IV, IVA and IVB)
p < 0.01 was considered statistically significant and shown in bold.

The heatmaps of subtype-specific genes were given in Figure 4. Overall, prognostic gene expression levels are in agreement with the DSS times. For instance, the patients with highly expressed CXCL10 were classified as high-risk group; otherwise, they are classified as low-risk groups. Even though there are a few patients who have high DSS time but were classified as a high-risk group, patients in the low-risk group have longer DSS time than the patients in the high-risk group.

**Figure 4.**
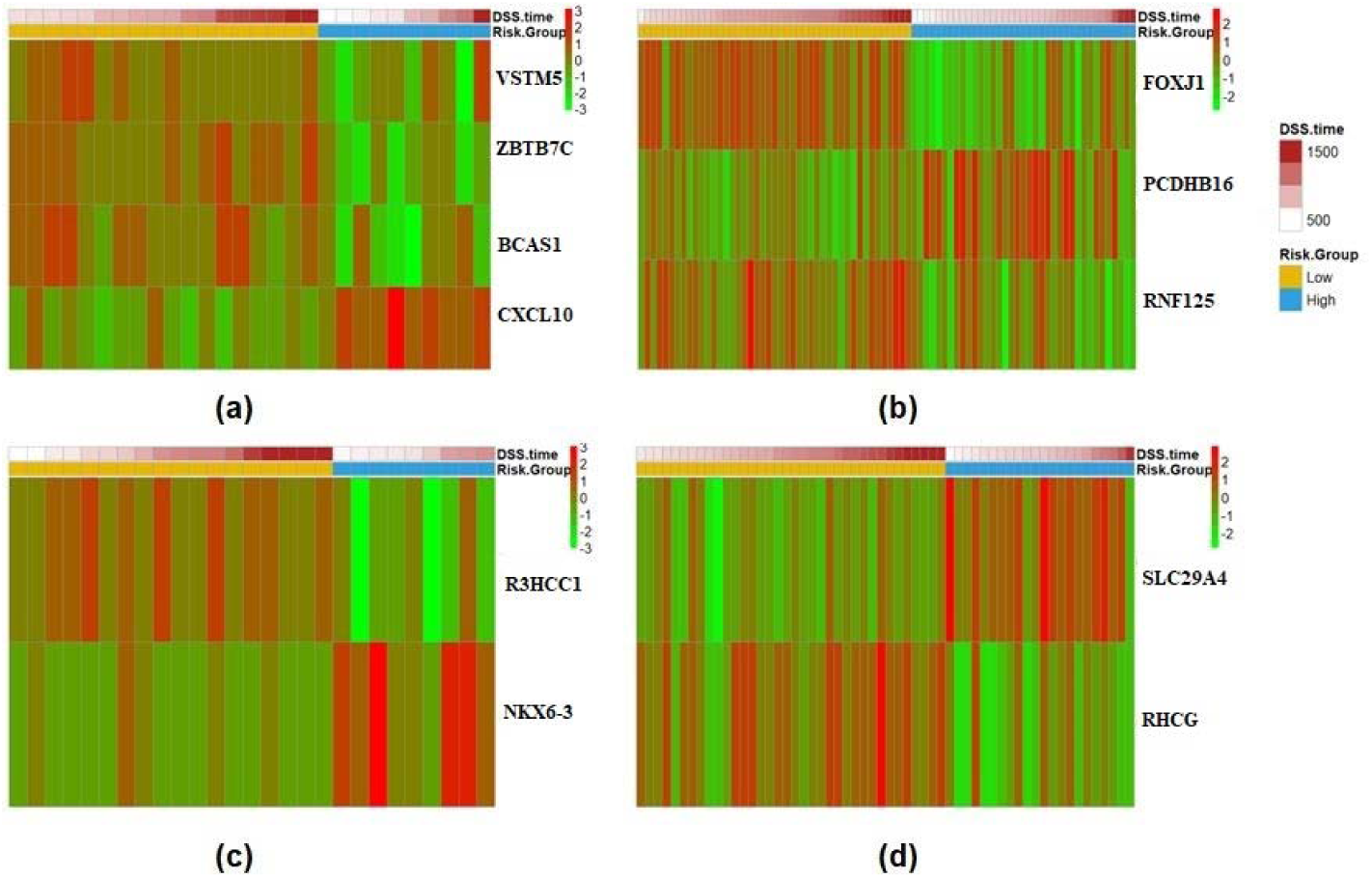
Heatmaps. Heatmaps representing the gene expression levels of subtype-specific markers in corresponding subtypes: (a) CMS1, (b) CMS2, (c) CMS3 and (d) CMS4. Rows represent the genes and columns represent the samples ordered according to the DSS time. Low-risk and high-risk group patients were represented with corresponding colored bars below the DSS time bar.

To validate the results, we merged TCGA COAD (260 patients) and Marshfield (35) cohorts to form the validation cohort. The validation results were shown in Figure 5 and Figure 6. The KM survival analysis indicated that the patients with high-risk scores in the CMS1, CMS2, and CMS4 subtypes had significantly poorer prognosis, compared to those with low-risk scores, as seen in the discovery cohort (Figure 5, p<0.05). Patients were divided into two groups, a low-risk group (146 patients) and a high-risk group (149 patients) by using 11 signature genes identified in this study. Furthermore, the KM analysis with log-rank test showed that our risk score approach was able to distinguish the low-risk group from the high-risk group using these subtype-specific signature genes (Figure 6, p<0.001).

**Figure 5.**
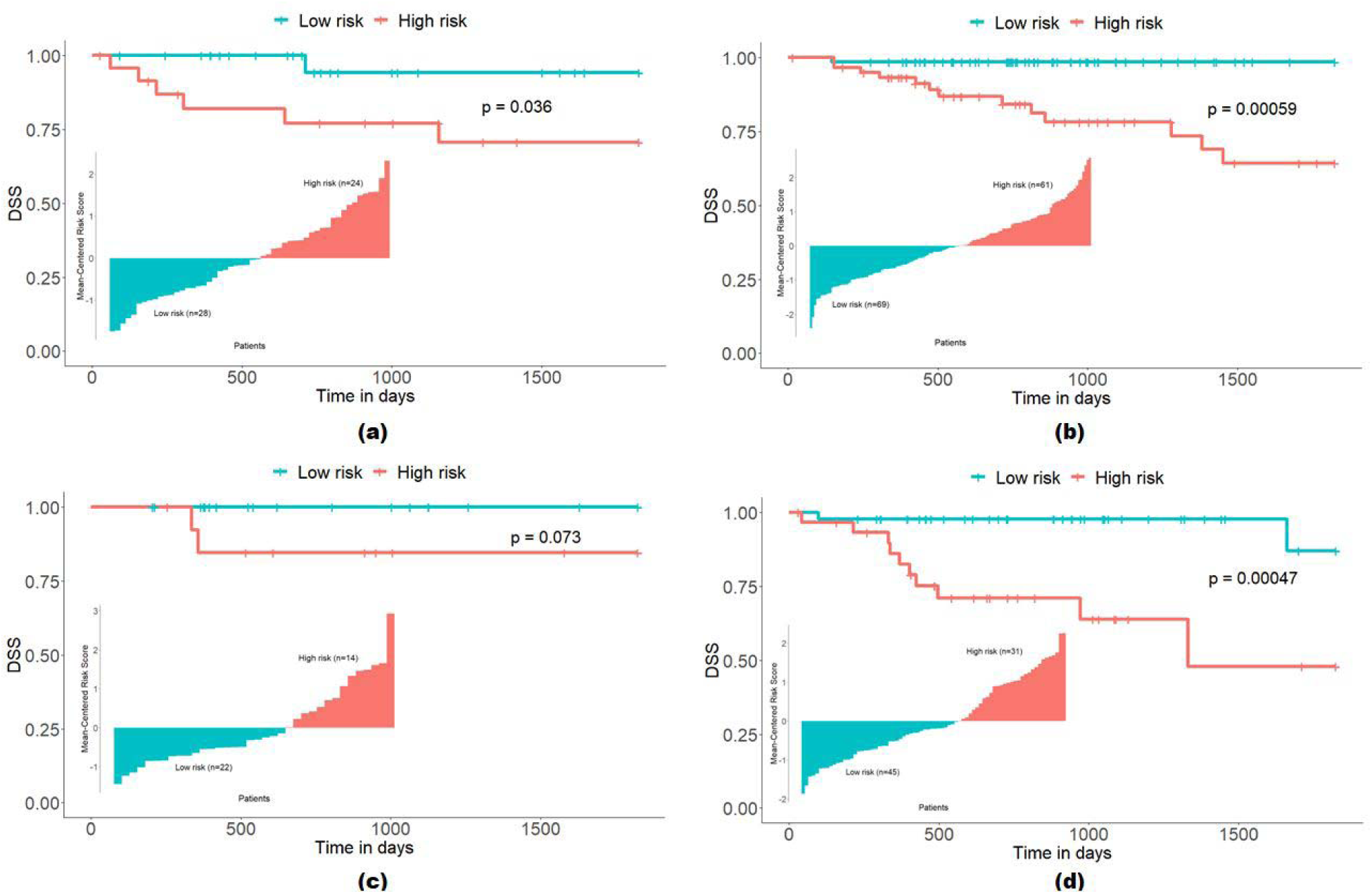
Patient stratification based on subtype risk scores with CMS specific prognostic markers in the validation cohort. Bar charts and KM survival curves for CSM1 (a), CMS2 (b), CMS3 (c) and CMS4 (d). Risk scores are given in a bar chart, where each bar indicates the subtype risk score for an individual colon cancer patient. Kaplan-Meier curve showing colon cancer patients with high and low-risk groups.

**Figure 6.**
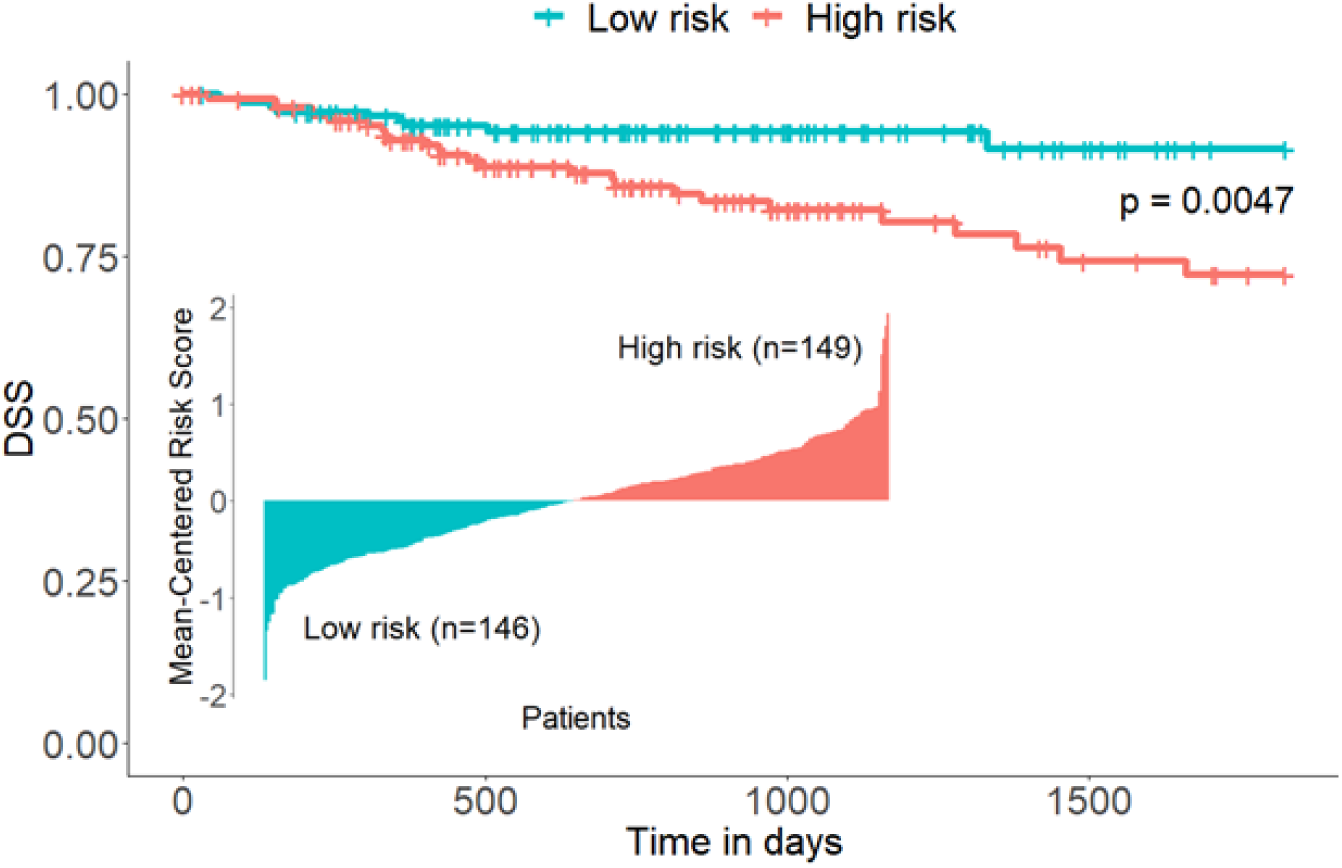
CRC patient stratification with 11 prognostic markers in the validation cohort. Risk scores are given in a bar chart, where each bar indicates the final risk score for an individual colon cancer patient. Kaplan-Meier curve showing colon cancer patients with high and low-risk groups. Subtype-specific signature genes can distinguish high-risk group from low-risk group with the log-rank *p*-value of 0.0047. Survival times are in days.

## DISCUSSION

With recent advancements in high-throughput technology providing data on colon cancer, a consensus by an international consortium classified colon cancer patients into four different subtypes using transcriptomic data ^13^. Although Bramsen et al. (2017) showed that subtype-specific genes can be used for prognosis prediction of colorectal cancer patients; there is still a gap in the literature in identifying the reliable subtype-specific prognostic genes. Therefore, in this study we developed a new fuzzy-based approach to identify consensus molecular subtype-specific genes for each CMS as FCM clustering is able to stratify patients using the natural structure of the data and handle the ambiguity in the data. Our approach has two key strengths: (i) instead of using recurrence free survival we used DSS data; and (ii) we used consensus molecular subtypes in order to identify subtype-specific genes.

Our study is specific to colon cancer RNA-seq data with additional CMS information and DSS data. Thus, we applied our approach to publicly available TCGA COAD data (77% of TCGA COAD) and validated the results with a larger cohort (combined TCGA COAD and Marshfield cohorts). These results could not be validated in CMS3 patients due to the low number of patients in the combined validation cohort. We suggest that it might be validated in a cohort with a higher CMS3 patient number. Results that are more accurate might be derived from a higher number of patients in cohorts; although current studies with RNA-Seq results of patients with clinical information such as CMS is very limited.

We observed that gene expression levels of prognostic genes are correlated with risk groups (Figure 4). For example, in immune response-associated CMS1, low expression of VSTM5, ZBTB7C and BCAS1 correlate with high-risk group, whereas high expression of CXCL10 correlates with the same risk group. There are no known tumor associated immune functions of the former genes, whereas CXCL10 has been previously identified as a prognostic marker of colon cancer with a well-known immunoregulatory role in multiple studies. Although no previous study identified CXCL10 as a CMS1-associated prognostic gene, our risk association of CXCL10 gene in colon cancer is supported with current literature^14–17^, further supporting the validity of our approach. Furthermore, this also provides the basis for the study of the unknown anti-tumor immune functions of our CMS1 associated prognostic genes in colon cancer models.

In CMS2, low expression of FOXJ1 and RNF125 correlate with high-risk group whereas high PCDHB16 expression belongs to the same risk group. FoxJ1 transcription factor is known to suppress APC^18^ and Wnt/β-catenin pathways^19^, whereas RNF125 is a protein ubiquitin ligase, which targets TP53^20^. Their roles in survival of CMS2 group of colon cancer patients is yet to be investigated.

While low expression of NKX6-3 correlates with low-risk group, low expression of R3HCC1 correlates with high-risk group in CMS3. NKX6-3 is a transcription factor with unknown associations in colon cancer. Although our study has the lowest number of patients in CMS3 group, disease association with prognostic markers could be studied with validation in larger cohorts.

Although high expression of SLC29A4 correlates with high-risk group, low expression of RHCG belongs to the same risk group in CMS4. SLC29A4 is a membrane protein with a described adenosine transport function^21^, which is yet to be associated with colon cancer survival.

There are also other studies, which focused on specific tumor stages such as ColoPrint^22^ for patients in early stage, ColDx^23^, ColoGuideEx^24^ for Stage II patients and ColoGuidePro^25^ for patients in Stage III. However, it should be noted that our study is stage independent but can be further investigated for identifying stage-specific genes. When we further grouped our validation cohort patients according to tumor stages, we observed that tumor stage III (Stage III, IIIA, IIIB and IIIIC combined) patients are best resolved according to prognosis with our CMS dependent genes (Supplementary Figure S2). We further examined the effect of these biomarkers in prognosis prediction with metastatic (Stage IV) and non-metastatic groups (Stage I, II, III combined). The 11 prognostic genes identified in this study can distinguish low-risk non-metastatic group from high-risk non-metastatic group whereas they cannot distinguish low-risk metastatic group from high-risk metastatic group (Supplementary Figure S3).

In this study, we developed a new approach in order to identify CMS specific signature genes that are predictive of survival and prognosis of colon adenocarcinoma patients. We have shown that these 11 signature genes which are colon adenocarcinoma related can be used for prognosis prediction of DSS based risk group of patients with colon adenocarcinoma. The 11 genes affecting survival are of great importance in terms of clinical decision support systems in predicting the prognosis of the disease and deciding on personalized adjuvant therapy, therefore they are potential candidates for mechanistic studies on colon cancer survival.

## SUMMARY TABLE

|  |  |
| --- | --- |
| **Problem** | Consensus Molecular Subtype (CMS)-based colon cancer classification lacks subtype-specific prognostic risk prediction with high confidence. |
| **What was already known on the topic** | • The absence of targeted therapy for primary colorectal cancer suggested the value of tumor stratification with CMSs as a prognostic approach (10).<br>• Identification of subtype-specific genes is proposed to improve the prediction of prognosis in colorectal cancer (9) |
| **What this study added to our knowledge** | • This study showed that an overall 11 CMS-specific genes is sufficient for efficient prognostic risk prediction and classification based on the transcriptomic data and disease-specific survival of colon cancer patients.<br>• Tumor stage grouping of the validation cohort suggested the best prediction of prognosis in stage III patients. |

## Data Availability

All data produced in the present study are available upon reasonable request to the authors

https://portal.gdc.cancer.gov

https://www.ncbi.nlm.nih.gov/geo/

## ACKNOWLEDGEMENTS

We would like to thank Dr. Steven A. Buechler for providing clinical data of Marshfield cohort. We also thank Drs. Gökhan Karakülah and Yavuz Oktay for their helpful discussions during the initiation of the study. Authors would like to thank Mrs. Buse Andaç Aktas for her contributions towards the preparation of the graphical abstract.

## DATA AVAILABILITY

The datasets supporting the conclusions of this article are publicly available and can be downloaded from TCGA data portal (https://portal.gdc.cancer.gov) and NCBI data portal (https://www.ncbi.nlm.nih.gov/geo/) or by using TCGAbiolinks R package.

---

**Supplementary Figure S1.**
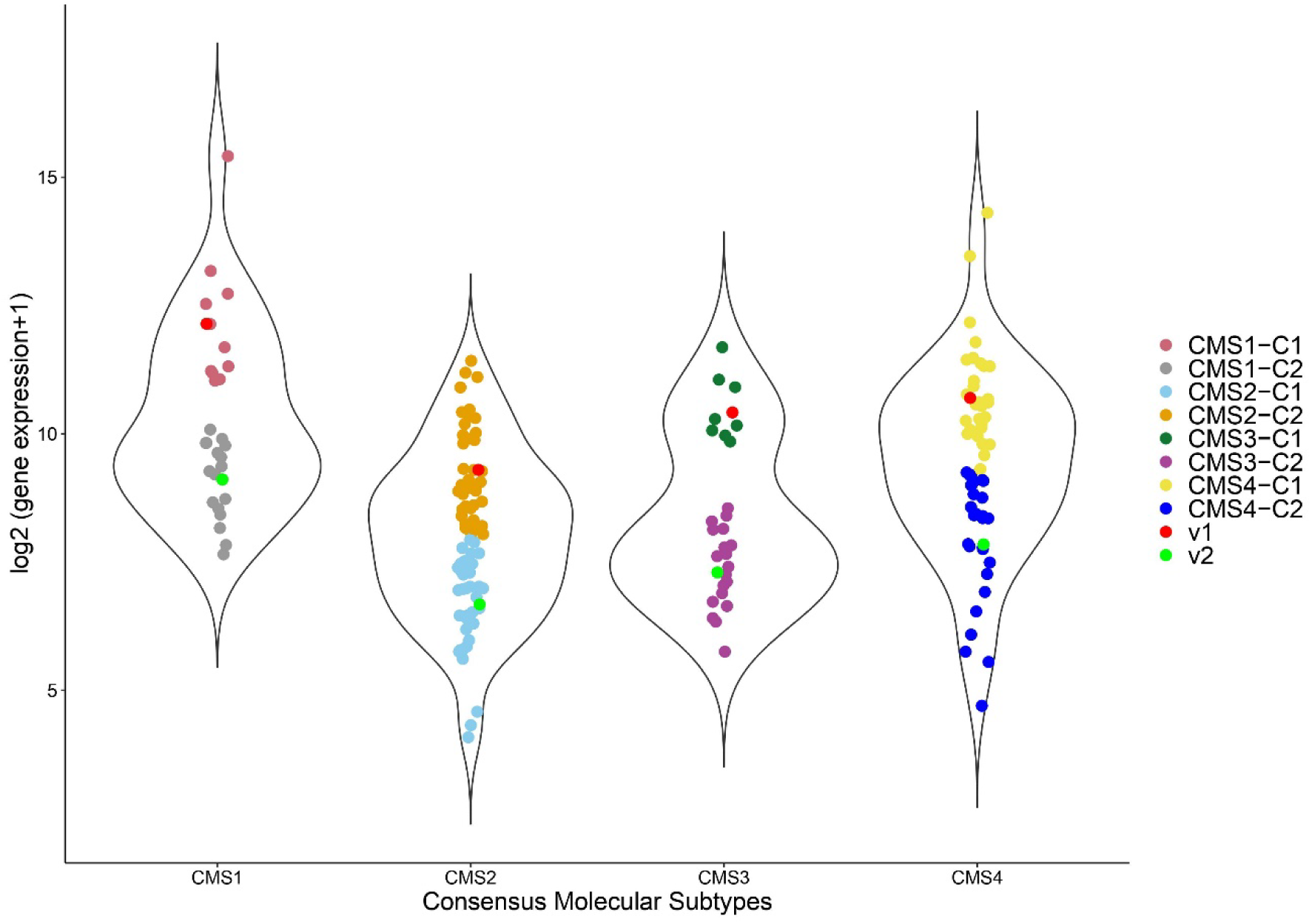
Representative violin plot showing FCM clustering for each CMS. Patients are stratified according to log2(CXCL10)+1 expression values into two clusters denoted by C1 and C2 for each consensus molecular subtype (CMS1-4), where v1 (red dot) and v2 (green dot) are cluster centers, respectively. Data points represent expression level of CXCL10 for each patient in each consensus molecular subtype (nCMS1=28, nCMS2=82, nCMS3=27, nCMS4=58).

**Supplementary Figure S2.**
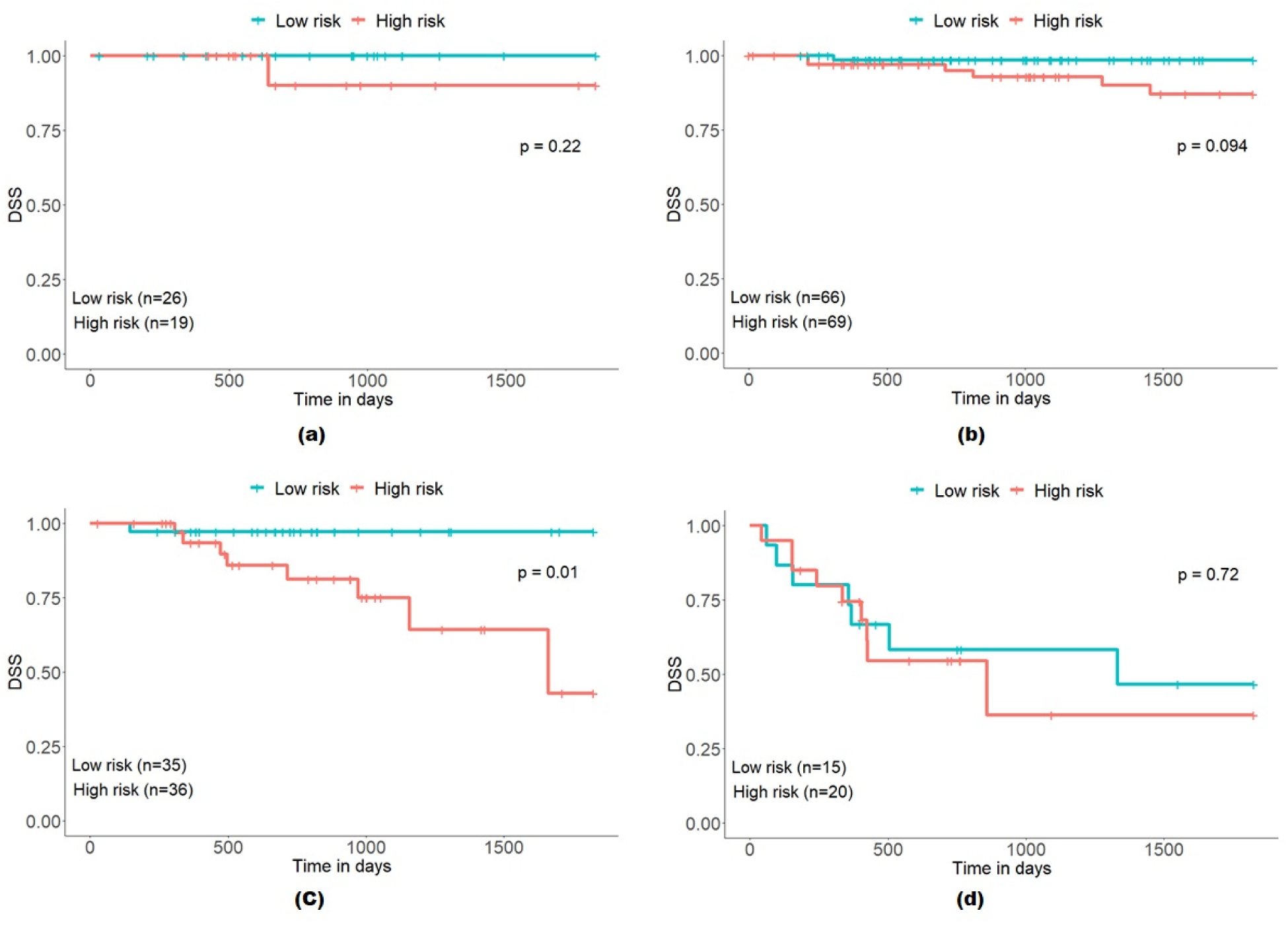
Patient stratification based on risk scores with 11 prognostic markers in the validation cohort. KM survival curves for Stage I (a), Stage II (b), Stage III (c) and Stage IV (d). Kaplan-Meier curves showing stage-specific colon cancer patients with high and low-risk groups.

**Supplementary Figure S3.**
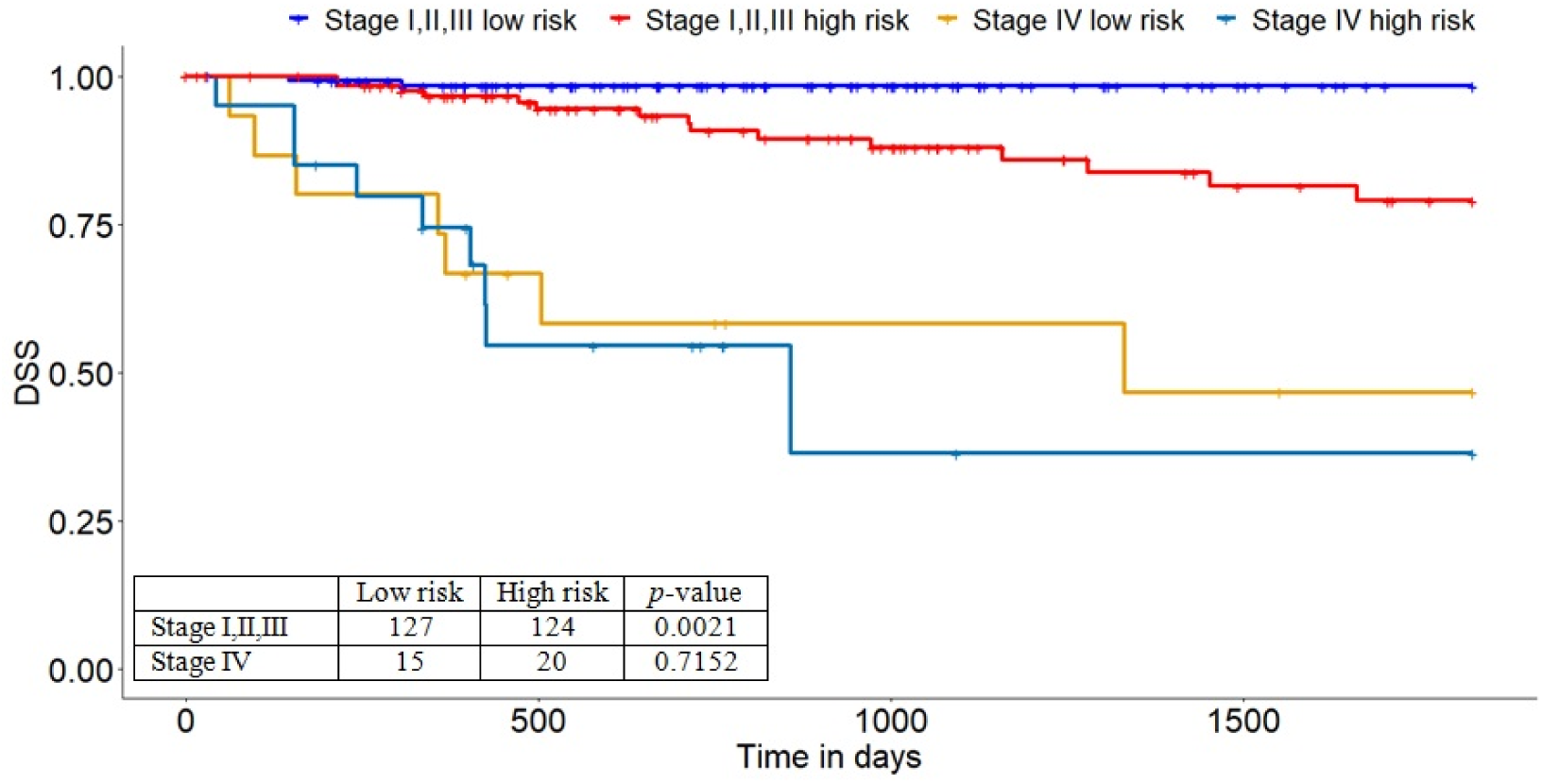
Patient stratification based on risk scores with 11 prognostic markers in the validation cohort. KM survival curves for Stage I, II, III (low and high-risk), and Stage IV (low and high-risk)

## REFERENCES

1 Guinney J, Dienstmann R, Wang X, De, et al. The consensus molecular subtypes of colorectal cancer. Nat Med. 2015; 21 (11): 1350–6.

2 Sung H, Ferlay J, Siegel RL, Laversanne M, et al. Global cancer statistics 2020: GLOBOCAN estimates of incidence and mortality worldwide for 36 cancers in 185 countries. CA Cancer J Clin. 2021; 71: 209–249. https://doi.org/10.3322/caac.21660.

3 Kuipers EJ, Grady WM, Lieberman D, et al. Colorectal cancer. Nat Rev Dis Primers. 2015; 1. https://doi.org/10.1038/nrdp.2015.65

4 Demirkol Canlı S, Seza EG, Sheraj I, et al. Evaluation of an aldo-keto reductase gene signature with prognostic significance in colon cancer via activation of epithelial to mesenchymal transition and the p70S6K pathway. Carcinogenesis. 2020 Sep 24; 41 (9): 1219–1228.

5 Gentles AJ, Newman AM, Liu CL, et al. The prognostic landscape of genes and infiltrating immune cells across human cancers. Nat Med. 2015 Aug; 21 (8): 938–945.

6 Kim SK, Kim SY, Kim CW, et al. A prognostic index based on an eleven gene signature to predict systemic recurrences in colorectal cancer. Exp Mol Med. 2019; 51: 1–12.

7 Liu J, Lichtenberg T, Hoadley KA, et al. An Integrated TCGA Pan-Cancer Clinical Data Resource to Drive High-Quality Survival Outcome Analytics. Cell. 2018 Apr 5; 173 (2): 400–416.

8 Sun D, Chen J, Liu L, et al. Establishment of a 12-gene expression signature to predict colon cancer prognosis. PeerJ. 2018 Jun 14; 6: e4942.

9 Bramsen JB, Rasmussen MH, Ongen H, et al. Molecular-Subtype-Specific Biomarkers Improve Prediction of Prognosis in Colorectal Cancer. Cell Rep. 2017 May; 19 (6): 1268–80.

10 Purcell R V., Schmeier S, Lau YC, et al. Molecular subtyping improves prognostication of Stage 2 colorectal cancer. BMC Cancer. 2019; 19 (1155).

11 Olsheski M, Schwartz D, Rineer J, et al. A Population-Based Comparison of Overall and Disease-Specific Survival Following Local Excision or Abdominoperineal Resection for Stage I Rectal Adenocarcinoma. J Gastrointest Cancer. 2013 Sep 6; 44 (3): 305–12.

12 Buechler SA, Stephens MT, Hummon AB, et al. ColoType: a forty gene signature for consensus molecular subtyping of colorectal cancer tumors using whole-genome assay or targeted RNA-sequencing. Sci Rep. 2020; 10 (1): 1–13.

13 Dienstmann R, Guinney J, Delorenzi M, et al. Colorectal Cancer Subtyping Consortium (CRCSC) Identifies Consensus of Molecular Subtypes. Ann Oncol. 2014; 25.

14 Zeng YJ, Lai W, Wu H, Liu L, Xu HY, Wang J, Chu ZH. Neuroendocrine-like cells -derived CXCL10 and CXCL11 induce the infiltration of tumor-associated macrophage leading to the poor prognosis of colorectal cancer. Oncotarget. 2016 May 10; 7 (19): 27394–407.

15 Zhao J, Wang Y, Gao J, Wang Y, Zhong X, Wu X, Li H. A nine-gene signature to improve prognosis prediction of colon carcinoma. Cell Cycle. 2021 May; 20 (10): 1021–1032.

16 Lu J, Chen Q. Transcriptome-based identification of molecular markers related to the development and prognosis of Colon cancer. Nucleosides Nucleotides Nucleic Acids. 2021; 40 (11): 1114–1124.

17 Song W, Yin H, Han C, Mao Q, Tang J, Ji Z, Yan X, Wang L, Liu S, Ai C. The role of CXCL10 in prognosis of patients with colon cancer and tumor microenvironment remodeling. Medicine (Baltimore). 2021 Sep 24; 100 (38): e27224.

18 Liu, K., Fan, J., Wu, J. 2017. “Forkhead box protein J1 (FOXJ1) is overexpressed in colorectal cancer and promotes nuclear translocation of β-catenin in SW620 cells”. Medical Science Monitor, 23, 856–866.

19 Caron, A., Xu, X., Lin, X. 2012. “Wnt/β-catenin signaling directly regulates Foxj1 expression and ciliogenesis in zebrafish Kupffer’s vesicle”. Development, 139 (3), 514–524.

20 Yang, L., Zhou, B., Li, X., Lu, Z., Li, W., Huo, X., & Miao, Z. (2015). RNF125 is a Ubiquitin-Protein Ligase that Promotes p53 Degradation. Cellular Physiology and Biochemistry, 35 (1), 237–245.

21 Engel K, Zhou M, Wang J. Identification and characterization of a novel monoamine transporter in the human brain. J Biol Chem 2004;279:50042–9

22 Tan IB, Tan P. An 18-gene signature (ColoPrint) for colon cancer prognosis. Nat Rev Clin Oncol. 2011 Mar 8; 8 (3): 131–3.

23 Felton J, Raufman J-P. Is the ColDx assay a valid prognostic marker for stage II colon cancer? Transl Cancer Res. 2016 Nov; 5 (S6): S1157–9.

24 Ågesen TH, Sveen A, Merok MA, et al. ColoGuideEx: a robust gene classifier specific for stage II colorectal cancer prognosis. Gut. 2012 Nov; 61 (11): 1560–7.

25 Sveen A, Agesen TH, Nesbakken A. ColoGuidePro[: A prognostic 7-gene expression signature for stage III colorectal cancer patients. Clin Cancer Res. 2012 Nov 1; 18 (21): 6001–10.

